# Relations Between Prenatal Sleep Health and Maternal Weight Retention 2 to 7 Years After a First Birth: The NuMoM2b-HHS

**DOI:** 10.64898/2026.08.23.26361117

**Authors:** Marquis. S. Hawkins, Rya B Clifton, Michele D. Levine, Namhyun Kim, Christina Personette, Mattina A. Davenport, Andrea C. Kozai, Rachel Kolko Conlon, David Phan, William Grobman, Jenna T. Ryan, Angela C. Ranzini, Jessica M. Page, David M. Haas, C. Noel Bairey Merz, George Saade, Lynn M. Yee, Phyllis C. Zee, Judith H. Chung, Janet Catov

## Abstract

**Background:** Poor prenatal sleep health is associated with greater gestational weight gain, but the contributions to longer-term maternal weight retention remain unclear.

**Purpose:** To examine associations between prenatal sleep health across multiple domains and maternal weight retention 2–7 years after a first birth.

**Methods:** Participants were from the nuMoM2b-Heart Health Study. Self-reported sleep was assessed during early (6-13 6/7 weeks) and mid-pregnancy (22–28 6/7 weeks) across six domains: regularity, quality, sleepiness, timing, efficiency, and duration. A multidimensional sleep health (MSH) score reflected the number of domains meeting healthy thresholds. Outcomes included maternal weight retention, total and substantial (≥11 lbs.), from pre-pregnancy to 2–7 years after a first birth. Associations were estimated using adjusted linear regression for total weight retention and Poisson regression with robust variance for substantial weight retention.

**Results:** The sample included 3,661 individuals with data in early (n = 2,962) and mid-pregnancy (n = 3,210). In early pregnancy, healthy sleep duration was associated with lower total weight retention, whereas healthy sleep regularity was unexpectedly associated with greater retention. In contrast, during mid-pregnancy, a higher MSH score was associated with a lower risk of substantial weight retention (RR = 0.96, 95% CI: 0.93–0.99). Healthy sleep duration and quality were the two individual domains associated with lower weight retention (2–3 lbs.).

**Conclusions:** In a prospective cohort of pregnant nulliparous individuals, healthy prenatal sleep, particularly during mid-pregnancy, was associated with less maternal weight retention 2-7 years after delivery. Future studies should estimate the causal effects of sleep health on long-term maternal weight retention.

## Introduction

Postpartum weight retention (PPWR) is a major public health concern with lasting implications for maternal cardiometabolic health.[1] Retaining weight after pregnancy increases the likelihood of entering subsequent pregnancies with a higher pre-pregnancy body mass index (BMI), which is associated with elevated risk of adverse pregnancy outcomes (e.g., preeclampsia, gestational diabetes), adverse neonatal outcomes (e.g., preterm birth), and long-term chronic disease, including type 2 diabetes, hypertension, and cardiovascular disease. [2, 3] Although gestational weight gain (GWG) and pre-pregnancy BMI have been consistently associated with PPWR [2–8], these factors alone don’t fully account for long-term weight retention. There is a need to identify additional modifiable behavioral predictors of PPWR that could be targeted during pregnancy.

Multidimensional sleep health is a modifiable behavior associated with GWG and obesity but has received comparatively little attention in the context of long-term maternal weight outcomes. Emerging research highlights the importance of conceptualizing sleep as a multidimensional health behavior that extends beyond sleep duration and quality. Dimensions such as sleep regularity, daytime sleepiness, timing, and efficiency remain relatively understudied in relation to postpartum weight outcomes, despite their established associations with obesity in general adult populations.[9, 10] Recent findings from the Nulliparous Pregnancy Outcomes Study: Monitoring Mothers-to-Be (nuMoM2b) cohort suggest that multidimensional sleep health (MSH), which reflects sleep patterns across multiple domains, is associated with gestational weight gain and may capture aspects of sleep behavior not reflected in any single characteristic.[11] However, little is known about whether individual sleep domains or MSH during pregnancy are associated with long-term maternal weight retention.

Racial and ethnic disparities in sleep health are also well documented in the general population and during pregnancy, and are rooted in structural and social inequities (e.g., residential segregation, neighborhood disadvantage) and social stressors (e.g., discrimination).[12–14] Black and Hispanic pregnant individuals experience higher rates of sleep disturbance and, in some cases, stronger physiological responses to sleep disruption, including heightened inflammation.[15–17] Evidence also suggests that associations between sleep and obesity may be stronger among Black adults compared with White adults.[17] However, few studies have examined whether prenatal sleep health is associated with long-term maternal weight outcomes differently across racial and ethnic groups.

To address these gaps, this study aimed to examine associations between prenatal sleep health—both individual domains and MSH—and maternal weight retention 2 to 7 years after birth, and to determine whether these associations differ by race and ethnicity. We hypothesized that poorer prenatal sleep health, indexed both by individual domains and the MSH score, would be associated with greater long-term maternal weight retention. Also, we hypothesized that associations between prenatal sleep health and long-term weight outcomes would differ by race and ethnicity, with stronger associations observed among Non-Hispanic Black participants, consistent with prior literature on sleep-related obesity disparities.[17]

## Methods

### Study Design and Population

This secondary analysis used data from the nuMoM2b Heart Health Study (nuMoM2b- HHS), a prospective cohort of pregnant nulliparous individuals designed to investigate the development of cardiovascular risk following a first pregnancy.[18] NuMoM2b-HHS enrolled participants from the original nuMoM2b cohort, which recruited 10,038 nulliparous individuals from eight ethnically diverse U.S. clinical centers between 2010 and 2013. Participants in the original nuMoM2b study were enrolled in the first trimester (visit 1, 6-13 6/7 weeks’ gestation) and followed them through delivery with standardized assessments conducted at up to three further visits: visit 2 (16-21 weeks’ gestation), visit 3 (22-28 6/7 weeks’ gestation), and visit 4 (delivery). These visits included structured interviews, self-administered questionnaires, anthropometric measurements, and biospecimen collection.[18, 19] The self-reported sleep data (primary exposure) was collected at visits 1 and 3. Hereafter, we refer to visit 1 as early pregnancy and visit 3 as mid-pregnancy.

All nuMoM2b participants were eligible for nuMoM2b-HHS if they had delivery information available, consented to future contact, were not currently pregnant, and were at least six months postpartum from any subsequent birth.[20] The in-person follow-up visit, attended by 4,508 participants, occurred 2 to 7 years (mean ∼ 3.02 years) after delivery and included clinical assessments, anthropometric measures, and questionnaires on lifestyle behaviors, psychosocial factors, and cardiometabolic health. For this secondary analysis, we included individuals who had data on the primary sleep health domains and key covariates (described further below) in early or mid-pregnancy, as well as weight data at the postpartum visit (**Supplemental Figure 1**). The Institutional Review Boards at all participating institutions approved this study, and all participants provided written informed consent prior to participation. All study procedures were conducted in accordance with the ethical standards of the responsible institutional research committees and with the 1964 Declaration of Helsinki and its later amendments.

### Exposure: sleep health

Sleep health was assessed at early and mid-pregnancy visits using a validated self-administered questionnaire.[18] The questionnaire evaluated multiple dimensions of sleep health based on participants’ experiences over the previous four weeks.

*Sleep timing* was defined as the midpoint between sleep onset and wake time (i.e., the sleep midpoint) and was averaged for both weekdays and weekends. [21] *Sleep regularity* was operationalized as the absolute difference between weekday and weekend sleep midpoints. *Sleep quality* was assessed using a series of items adapted from the Women’s Health Initiative Insomnia Rating Scale.[22] Participants also rated the overall quality of their typical night’s sleep in the past 4 weeks on a 5-point scale: very sound or restful, sound or restful, quality, restless, or very restless. *Daytime sleepiness* was assessed using the Epworth Sleepiness Scale (ESS) , which measures the likelihood of dozing off in various routine situations (e.g., while reading, watching television, riding in a car).[23] Responses were summed to generate a total score ranging from 0 to 24, with higher scores indicating greater sleep propensity. [23] The *sleep efficiency* domain was operationalized as wake after sleep onset (minutes spent awake during the night). *Sleep duration* was estimated as the weighted average of weekday and weekend self-reported typical number of hours slept per night.

Healthy sleep in each domain was defined using the National Sleep Health Foundation recommendations or empirical data when recommendations were not available.[24, 25] In brief, healthy sleep was defined as regular sleep timing (<1-hour weekday–weekend difference), adequate duration (7–9 hours/night), good subjective sleep quality, not sleepy (ESS score <10), a sleep midpoint between 2:00 and 4:00 AM, and limited wake after sleep onset (<40 minutes). To calculate *MSH*, we created a simple composite score indicating the number of “healthy” sleep domains. MSH scores ranged from 0 to 6; higher values indicating better MSH.[26]

### Outcome: long-term maternal weight retention

The primary outcomes were total and substantial maternal weight retention from pre-pregnancy to 2–7 years after childbirth. At the nuMoM2b-HHS follow-up visit, participants underwent standardized anthropometric measurements performed by trained study staff using a uniform protocol across sites.[18] Total maternal weight retention was calculated as the difference in weight (in pounds) between the participant’s pre-pregnancy weight (determined at Visit 1 based on self-report) and measured weight at follow-up visit. Substantial weight retention was defined as ≥11 lbs. relative to pre-pregnancy weight, a threshold supported by prior research linking this level of postpartum weight retention to increased cardiometabolic risk.[1, 27]

### Demographic characteristics and covariates

A comprehensive set of covariates was also included across sociodemographic, health behavior, and clinical domains. Covariates included in the adjusted models were age, race/ethnicity, education level, marital status, government insurance status, smoking status, and pre-pregnancy BMI. Physical activity and depressive symptoms are reported as descriptive characteristics only and were not included as model covariates. Sociodemographic characteristics were self-reported using standardized surveys. Race/ethnicity was self-reported by participants. Participants self-reported smoking status three months before pregnancy. Physical activity was estimated using modified questions for the Behavioral Risk Factor Surveillance System questionnaire.[28] Depressive symptoms were assessed using the Edinburgh Postnatal Depression Scale (EPDS), a 10-item self-report measure widely used in perinatal populations with demonstrated reliability and validity.[29]

### Statistical Analysis

All data preparation, statistical modeling, and visualization were conducted using R (version 4.5.0). We used the gtsummary package to generate tables for descriptive characteristics and primary outcomes.[30] Analyses were conducted using model-specific casewise deletion (i.e., participants missing the outcome, exposure, or covariates for a given model were excluded). This casewise deletion process was used when examining associations in the overall sample and when conducting race-stratified analyses. Descriptive statistics were calculated for all variables. Continuous variables were summarized using means (standard deviations) or medians (interquartile ranges). Categorical variables were summarized with frequencies and percentages. Statistical significance was defined as a two-sided p-value < .05. Given that sleep domains were specified a priori based on MSH frameworks and are conceptually and empirically correlated, we did not adjust for multiple comparisons; findings should therefore be interpreted in the context of the overall pattern of results.

We examined associations between early- and mid-pregnancy prenatal sleep health—individual domains and MSH scores—and the two postpartum weight outcomes, modeling each exposure separately. We used linear regression models to examine associations with total weight retention. We used Poisson regression models with a log link and robust standard errors (HC3 sandwich estimator) to examine associations with substantial weight retention. All models were adjusted for covariates associated with prenatal sleep and weight, including age, race/ethnicity, education, marital status, government insurance status, pre-pregnancy BMI, and smoking status. Beta-coefficients and relative risks (RR) and 95% confidence intervals (CI) were presented for total and substantial weight retention respectively.

We included sleep-race interaction terms in each model to assess whether associations differed across racial/ethnic groups. Non-Hispanic White, Non-Hispanic Black, and Hispanic groups were included in the race-stratified analyses, while Asian and individuals who did not identify according to the aforementioned categories were excluded from stratification due to their small sample sizes. We performed stratified analysis when sleep-race interaction terms were statistically significant (p<0.05).

For models of substantial weight retention, we examined whether the association between prenatal sleep health and weight retention differed across racial and ethnic groups on the additive scale by calculating the Relative Excess Risk due to Interaction (RERI). We focused on additive interaction because it reflects whether the absolute burden of substantial weight retention among those exposed to both unhealthy sleep and a minoritized racial/ethnic identity exceeds what would be expected from each association acting independently — a quantity relevant for assessing where intervention might shift absolute risk within a given subgroup. Consistent with our framing of race and ethnicity as markers of structural and social context rather than biologic risk factors, RERI here characterizes how the magnitude of the sleep–weight association on the absolute-risk scale varies across racial and ethnic groups. RERI was calculated as RERI = RR□□− RR□□− RR□□+ 1, with each relative risk referenced to Non-Hispanic White participants with healthy sleep on the domain being tested. The first subscript indexed sleep (1 = unhealthy, 0 = healthy on that domain) and the second indexed race and ethnicity (1 = Non-Hispanic Black or Hispanic, 0 = Non-Hispanic White). RR reflected the risk associated with unhealthy sleep among Non-Hispanic White participants, RR reflected the risk associated with being Non-Hispanic Black or Hispanic among those with healthy sleep, and RR reflected the risk among those with both unhealthy sleep and minoritized racial or ethnic identity.

A RERI of zero is consistent with the two associations combining additively, with no departure from additivity. A positive RERI is consistent with positive additive interaction (“synergy”), in which the absolute increase in risk associated with unhealthy sleep is greater among the minoritized racial or ethnic group than among Non-Hispanic White participants. A negative RERI is consistent with negative additive interaction (“antagonism”), in which the absolute increase in risk associated with unhealthy sleep is smaller in the minoritized group than in Non-Hispanic White participants. As an approximate guide to magnitude, an RERI of 0.30 indicates a joint relative risk that is 0.30 units higher than expected if the two associations operated independently. Confidence intervals were calculated using the delta method, with 95% intervals excluding zero interpreted as a statistically significant departure from additivity.

## Results

Of the 4,508 participants who attended the in-person nuMoM2b-HHS visit, 3,661 met eligibility criteria for early- (n = 2,962) or mid-pregnancy (n = 3,210) analytic samples, respectively (**see Supplemental Figure 1**). Compared with excluded participants, those included in the analytic samples were generally older, were more likely to be Non-Hispanic White, had higher educational attainment and income, and were less likely to have government insurance or to report smoking prior to pregnancy (**Supplemental Table 1**). Although differences in pre-pregnancy BMI and early pregnancy MSH scores were statistically significant, absolute differences were modest.

Included participants had a median baseline age of 28.0 (IQR: 23.0, 31.0 years) and a median pre-pregnancy BMI of 23.9 (IQR: 21.4, 28.3 kg/m^2^). 67.8% of participants were Non-Hispanic White, 10.9% were Non-Hispanic Black, 13.5% were Hispanic, 3.3% were Asian, and 4.4% identified as belonging to other races. Most participants were educated beyond high school, married, had incomes substantially above the federal poverty level, and had commercial insurance. 15.1% smoked during the three months before pregnancy (**Table 1**). Sleep health characteristics of included participants at early and mid-pregnancy are described in **Supplemental Table 2**.

**Table 1.**
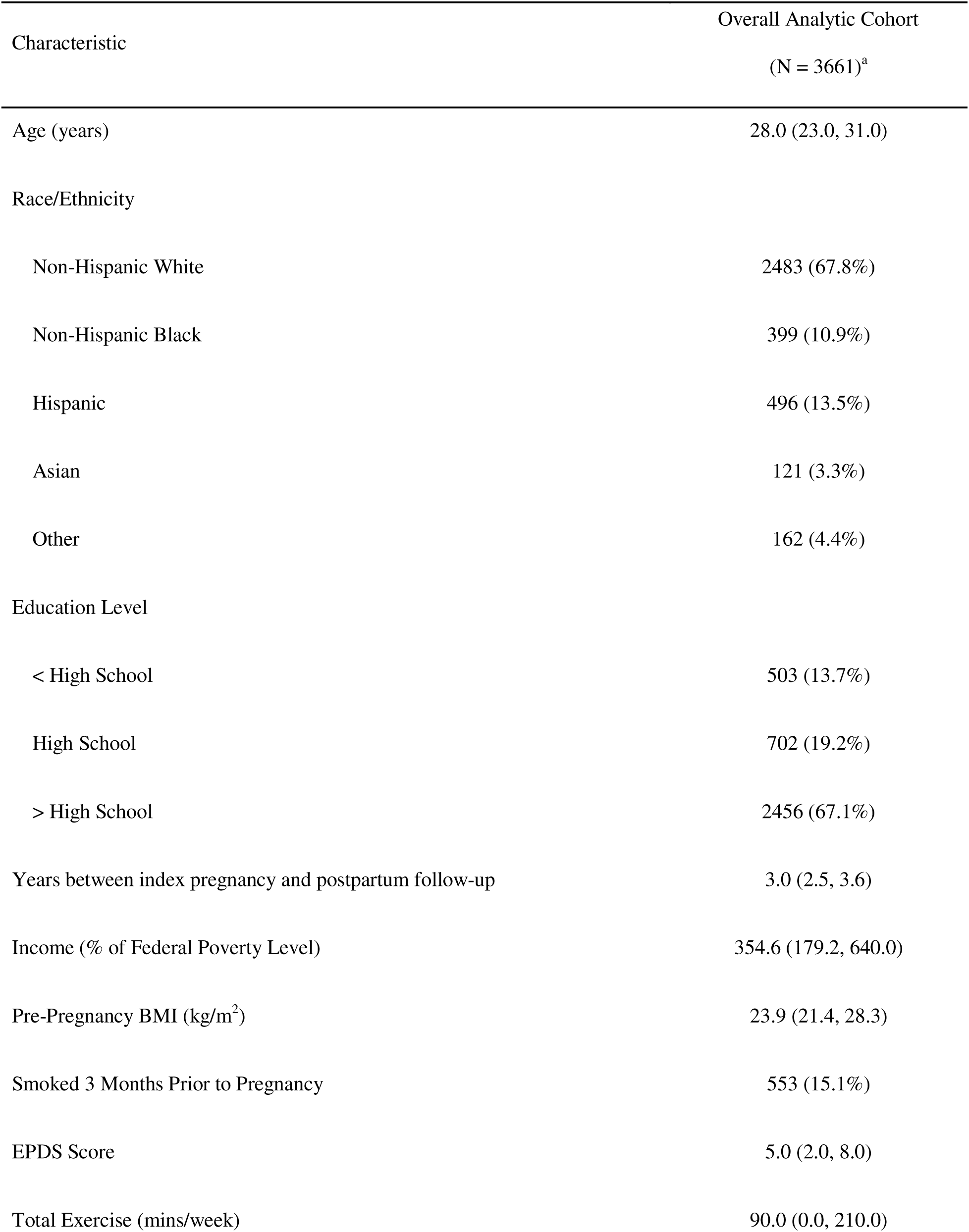
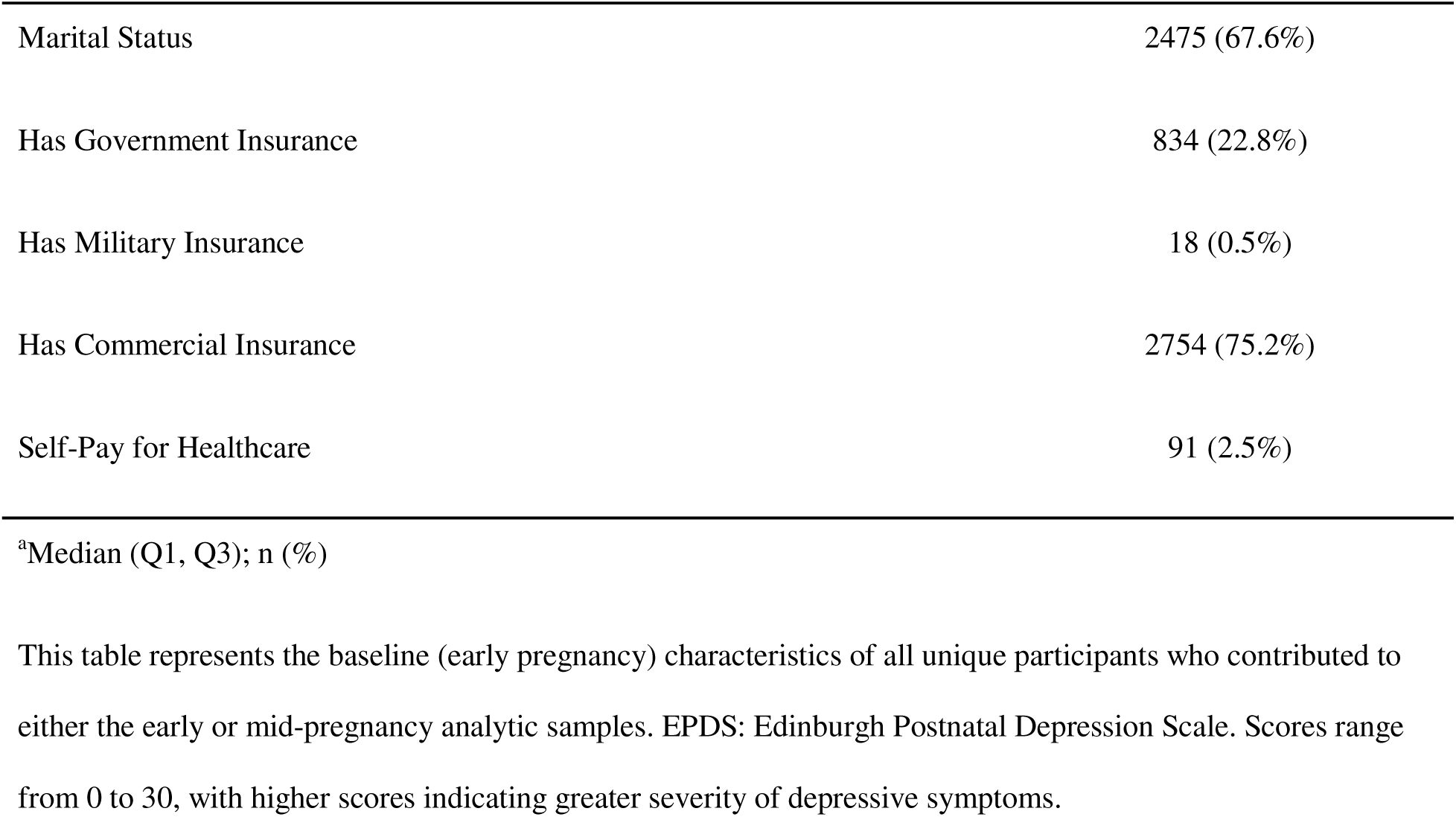
Baseline Characteristics of the Analytic Cohort.

### Continuous weight retention

In early pregnancy, associations between sleep health and continuous weight retention were inconsistent (**Table 2**). The MSH score was not significantly associated with continuous weight retention (β = –0.60, 95% CI: – 1.34, 0.14). Among individual domains, participants with healthy sleep duration (7–9 hours/night) retained 2.1 fewer lbs. compared to those sleeping <7 or >9 hours (95% CI: –4.0, –0.26). Conversely, participants with healthy sleep regularity retained 1.9 more lbs. than those with less regular sleep (95% CI: 0.05, 3.70). Other domains did not show statistically significant associations with continuous weight retention. Sleep–race interaction terms were not statistically significant for any early pregnancy domain (**Supplemental Table 3**).

**Table 2.** Associations between pregnancy sleep health indicators and continuous postpartum weight retention (lbs.).

| Sleep Domain | Early Pregnancy<br>N = 2962 |  | Mid-Pregnancy<br>N = 3210 |  |
| --- | --- | --- | --- | --- |
|  | Beta | 95% CI<sup>a</sup> | Beta | 95% CI<sup>a</sup> |
| Multidimensional Sleep Health | -0.60 | -1.34, 0.14 | -0.82 | -1.47, -0.17 |
| Sleep Timing |  |  |  |  |
| Outside of 2-4 AM window | — | — | — | — |
| Between 2-4 AM | -0.65 | -2.64, 1.34 | -0.89 | -2.71, 0.92 |
| Sleep Duration |  |  |  |  |
| < 7 or > 9 hours | — | — | — | — |
| 7-9 hours | -2.1 | -4.00, -0.26 | -2.4 | -4.12, -0.70 |
| Sleep Regularity |  |  |  |  |
| ≥ 1 hour | — | — | — | — |
| < 1 hour | 1.9 | 0.05, 3.70 | 0.98 | -0.64, 2.60 |
| Wake After Sleep Onset |  |  |  |  |
| > 40 minutes | — | — | — | — |
| ≤ 40 minutes | -0.62 | -2.92, 1.68 | -0.93 | -2.88, 1.02 |
| Sleep Quality |  |  |  |  |

| Sleep Domain | Early Pregnancy<br>N = 2962 |  | Mid-Pregnancy<br>N = 3210 |  |
| --- | --- | --- | --- | --- |
|  | Beta | 95% CI<sup>a</sup> | Beta | 95% CI<sup>a</sup> |
| Self-reported as worse than average | — | — | — | — |
| Self-reported as average or better | -1.5 | -3.73, 0.72 | -2.4 | -4.31, -0.44 |
| Daytime Sleepiness |  |  |  |  |
| Excessive Sleepiness ($\geq 10$ points) | — | — | — | — |
| Normal ($< 10$ points) | -1.6 | -3.45, 0.26 | -1.1 | -3.02, 0.78 |

<sup>a</sup>All models adjusted for age, race, education, marital status, pre-pregnancy BMI, and smoking status.

In mid-pregnancy, higher MSH scores and healthier sleep in several individual domains—particularly duration and quality—were associated with lower continuous weight retention in the overall sample (**Table 2**). Specifically, each additional point in the MSH score was associated with 0.82 lbs. less retained weight (95% CI: – 1.47, –0.17). Among the individual domains, those with a healthy sleep duration had 2.4 lb. less weight retention (95% CI: –4.12, –0.70) than those who did not. Healthy sleep quality was also associated with less retained weight (β = –2.4, 95% CI: –4.31, –0.44). Estimates varied by race (**Supplemental Table 3**). Specifically, we found a statistically significant interaction between race and mid-pregnancy sleep timing and sleep quality for Hispanic participants compared to Non-Hispanic White participants. The sleep timing × Hispanic interaction term was −9.38 lbs. (95% CI: −15.26, −3.51) relative to Non-Hispanic White participants. The sleep quality × Hispanic interaction term was 6.92 lbs. (95% CI: 0.64, 13.19) relative to Non-Hispanic White participants. In stratified models for these domains (**Supplemental Table 4**), Hispanic participants with a sleep midpoint within the 2:00–4:00 AM window retained more weight (β = 7.0, 95% CI: 1.34, 12.59), whereas those with healthy sleep quality retained less (β = –7.9, 95% CI: –14.59, –1.25). No other mid-pregnancy sleep–race interactions were statistically significant for continuous weight retention.

### Substantial weight retention (≥11 lbs.)

In early pregnancy (**Table 3**), most sleep domains showed small or non-statistically significant associations with substantial weight retention. There were statistically significant interactions between race and MSH and daytime sleepiness (**Supplemental Table 5**). Higher MSH scores (RR = 1.1, 95% CI: 1.02, 1.21) and normal daytime sleepiness (RR = 1.4, 95% CI: 1.09, 1.83) were each associated with a higher risk of substantial weight retention among Hispanic participants, a direction opposite to that observed among Non-Hispanic White and Non-Hispanic Black participants (**Supplemental Table 6**). No other early-pregnancy sleep–race interactions were statistically significant.

**Table 3.** Associations between pregnancy sleep health indicators and substantial postpartum weight retention (≥11 lbs.).

|  | Early Pregnancy<br>N = 2962 |  | Mid-Pregnancy<br>N = 3210 |  |
| --- | --- | --- | --- | --- |
| Sleep Domain | Risk Ratio | 95% CI<sup>a</sup> | Risk Ratio | 95% CI<sup>a</sup> |
| Multidimensional Sleep Health | 0.99 | 0.95, 1.02 | 0.96 | 0.93, 0.99 |
| Sleep Timing |  |  |  |  |
| Outside of 2-4 AM window | — | — | — | — |
| Between 2-4 AM | 0.99 | 0.90, 1.08 | 0.97 | 0.89, 1.06 |
| Sleep Duration |  |  |  |  |
| < 7 or > 9 hours | — | — | — | — |
| 7-9 hours | 0.97 | 0.88, 1.06 | 0.92 | 0.84, 1.00 |
| Sleep Regularity |  |  |  |  |
| $\geq 1$ hour | — | — | — | — |
| < 1 hour | 1.1 | 0.98, 1.17 | 1.0 | 0.96, 1.13 |
| Wake After Sleep Onset |  |  |  |  |
| > 40 minutes | — | — | — | — |
| $\leq 40$ minutes | 0.98 | 0.87, 1.09 | 0.93 | 0.85, 1.03 |
| Sleep Quality |  |  |  |  |
| Self-reported as worse than average | — | — | — | — |

Table 3 – Associations between pregnancy sleep health indicators and substantial postpartum weight retention ( $\geq 11$ lbs.).
| Sleep Domain | Early Pregnancy |  | Mid-Pregnancy |  |
| --- | --- | --- | --- | --- |
|  | N = 2962 |  | N = 3210 |  |
|  | Risk Ratio | 95% CI<sup>a</sup> | Risk Ratio | 95% CI<sup>a</sup> |
| Self-reported as average or better | 0.94 | 0.84, 1.04 | 0.88 | 0.80, 0.96 |
| Daytime Sleepiness |  |  |  |  |
| Excessive Sleepiness ($\geq 10$ points) | — | — | — | — |
| Normal ($< 10$ points) | 0.97 | 0.88, 1.06 | 0.91 | 0.83, 1.00 |

<sup>a</sup>All models adjusted for age, race, education, marital status, pre-pregnancy BMI, and smoking status.

During mid-pregnancy (**Table 3**), higher MSH scores were associated with a statistically lower risk of substantial weight retention in the overall sample (RR = 0.96, 95% CI: 0.93, 0.99). Among individual domains, healthy sleep quality was associated with lower risk (RR = 0.88, 95% CI: 0.80, 0.96); healthy sleep duration (RR = 0.92, 95% CI: 0.84, 1.00) and normal daytime sleepiness (RR = 0.91, 95% CI: 0.83, 1.00) showed estimates in the same direction, with confidence intervals that included the null. Other domains, including sleep timing, sleep regularity, and wake after sleep onset, showed small or non-significant associations in the overall sample. There were statistically significant interactions with sleep timing and quality among Hispanic participants (**Supplemental Table 5**). Specifically, Hispanic participants with a sleep midpoint within the 2:00–4:00 AM window had a higher risk of substantial weight retention (RR = 1.26, 95% CI: 1.01, 1.51). Sleep quality was associated with a lower risk of substantial weight retention among Hispanic participants (RR = 0.71, 95% CI: 0.59, 0.87) than among Non-Hispanic White participants. No other mid-pregnancy sleep–race interactions were statistically significant.

## Discussion

In this large prospective cohort, multidimensional sleep health during mid-pregnancy—rather than early pregnancy—was associated with maternal weight retention 2–7 years after birth. Healthy sleep duration, better subjective sleep quality, and higher multidimensional sleep health (MSH) scores during mid-gestation were each associated with lower retained weight and reduced risk of substantial weight retention. Each additional point on the MSH score corresponded to nearly one pound less retained weight and a 4% lower risk of retaining ≥11 lbs. (RR = 0.96, 95% CI: 0.93, 0.99), suggesting that cumulative sleep health may meaningfully be associated with long-term maternal weight trajectories. In contrast, associations in early pregnancy were inconsistent. Together, these findings extend prior research focused on postpartum sleep by demonstrating that sleep patterns established during pregnancy, particularly in mid-gestation, may have lasting implications for maternal weight regulation years beyond childbirth.

The associations observed in mid-pregnancy raise the possibility that mid-pregnancy sleep health is more closely linked to long-term weight outcomes than early-pregnancy sleep health, although this design cannot establish a sensitive window or any programming mechanism. Sleep disruption increases across gestation [31], coinciding with profound hormonal, metabolic, and inflammatory changes that influence insulin sensitivity, appetite regulation, and energy storage. Poor sleep duration and quality are known to alter glucose metabolism, appetite-regulating hormones, circadian alignment, and behavioral energy balance in non-pregnant adults.[32–34] During pregnancy, when metabolic demands are already heightened, sleep disruption may amplify physiologic stress in ways that persist beyond delivery.[35] Importantly, the MSH framework captures the cumulative and potentially synergistic effects of co-occurring sleep disturbances, rather than isolating single characteristics.[11, 26, 36] The observation that higher MSH was associated with lower long-term weight retention supports the conceptualization of sleep as an integrated behavioral and physiological construct that may influence maternal cardiometabolic trajectories across the reproductive life course. Individual effect sizes were modest. Because the MSH score is cumulative, participants meeting healthy criteria in more domains retained less weight on average; whether improving multiple domains would produce comparable reductions cannot be determined from these observational data. Weight retained after a first birth is associated with higher pre-pregnancy BMI in subsequent pregnancies, so small differences could plausibly accumulate across the reproductive life course; however, this study examined neither subsequent pregnancies nor cardiometabolic endpoints.

Prior studies have primarily focused on sleep during the early postpartum period, demonstrating that women sleeping fewer than five hours per night at 3 to 6 months postpartum are two to three times more likely to retain substantial weight one year after delivery.[3, 6, 27]. These findings established sleep as a behavioral factor related to postpartum weight retention. The present study extends this literature by shifting the window of exposure to pregnancy itself. Our results suggest that sleep patterns established during gestation, particularly in mid-pregnancy, may precede and potentially contribute to long-term maternal weight trajectories years after childbirth. Identifying prenatal sleep as an early marker of long-term weight risk broadens the temporal framework for considering behavioral interventions.

Overall, most sleep–race interaction terms were not statistically significant, and race-stratified patterns were generally similar in direction and magnitude across racial and ethnic groups. The clearest exceptions involved Hispanic participants: for continuous weight retention, significant interactions were observed for mid-pregnancy sleep timing and sleep quality; for substantial weight retention, significant interactions were observed for early-pregnancy MSH and daytime sleepiness and for mid-pregnancy sleep timing and sleep quality, all among Hispanic participants. Whether these findings reflect true differential associations or chance variation, given the number of comparisons tested, remains uncertain and warrants replication before definitive conclusions can be drawn. Pregnancy itself represents a period of heightened physiologic demand and has been described as a metabolic “stress test” for future cardiometabolic health [18, 35], and its metabolic adaptations may interact with pre-existing lived contexts—including occupational demands, caregiving responsibilities, environmental exposures, and chronic stress—in ways that shape how sleep disruption influences long-term weight regulation.[13, 37] Racial and ethnic disparities in sleep health during pregnancy are well documented [12, 38], and prior work suggests that sleep– obesity associations may differ across populations.[17] However, the largely null pattern of interaction results was contrary to our hypothesis, which was based on evidence that sleep–obesity associations differ by race among non-pregnant adults. These findings raise the possibility that prenatal sleep health does not differentially contribute to weight retention 2 to 7 years postpartum across racial and ethnic groups. Future studies should examine associations between postpartum multidimensional sleep health and weight by race, explore perinatal sleep trajectories as predictors of long-term weight outcomes, and incorporate combinations of subjective and objective sleep assessment tools to further interrogate these findings.

The finding that healthy sleep regularity in early pregnancy was associated with modestly greater weight retention warrants careful consideration, as it runs counter to our hypothesis and to prior evidence linking irregular sleep timing to metabolic dysregulation in general adult populations.[9] One potential explanation concerns the operationalization of sleep regularity in a pregnant sample. The healthy regularity threshold used here (i.e., less than one hour of weekday-to-weekend difference in sleep midpoint) was derived from frameworks developed in non-pregnant adults.[26] During early pregnancy, some individuals may intentionally extend weekend sleep in response to fatigue, nausea, or disrupted weeknight sleep, such that greater weekday-weekend variability reflects adaptive compensatory behavior rather than pathological circadian misalignment.[31] Under this interpretation, women with low variability may not need or be accessing the restorative benefit of weekend sleep extension. A second possibility is that highly regular sleep schedules during early pregnancy serve as a proxy for inflexible occupational or structural demands (e.g., fixed-shift work or demanding employment schedules) that constrain sleep timing while simultaneously limiting opportunities for physical activity and health-promoting behaviors.[13, 37] In this context, sleep regularity may capture social and structural constraints rather than circadian health. It is also notable that the regularity finding was present in early pregnancy but attenuated and non-significant in mid-pregnancy, and that the multidimensional sleep health composite, which integrates regularity alongside duration, quality, timing, efficiency, and alertness, was consistently associated with lower weight retention in the expected direction. This pattern suggests that when considered in isolation, regularity may not adequately capture the broader behavioral and metabolic significance of sleep as a multidimensional construct.[26, 36] Taken together, these findings underscore the need for future research to examine the specific dimensions of sleep regularity most relevant to pregnant populations, including the potential metabolic role of compensatory sleep and the structural determinants that constrain circadian flexibility during the perinatal period.

This study has several notable strengths that enhance its relevance to behavioral medicine. It leverages one of the largest socioeconomically, racially, and ethnically diverse U.S. pregnancy cohorts to examine multidimensional sleep health during pregnancy in relation to long-term maternal weight outcomes extending up to seven years postpartum. The prospective design, with repeated assessments of sleep in early and mid-gestation and standardized anthropometric follow-up years after delivery, allows for temporal ordering of sleep exposures and long-term weight trajectories. Importantly, the use of a multidimensional sleep health framework moves beyond single-domain metrics and captures the cumulative behavioral patterning of sleep across duration, quality, timing, regularity, efficiency, and daytime alertness. This integrated approach aligns with emerging scientific statements conceptualizing sleep as a modifiable, multidimensional health behavior with cardiometabolic implications. By situating prenatal sleep within a life-course framework of maternal cardiometabolic risk, this study extends sleep science into an earlier developmental window with translational potential for preventive intervention.

Several limitations should be considered when interpreting these findings. First, sleep was assessed using validated self-report instruments rather than objective measures such as actigraphy or polysomnography. Although subjective sleep reports may introduce measurement error and do not capture physiologic sleep architecture or circadian phase directly, perceived sleep experience is itself a core dimension of sleep health and has been shown to be strongly associated with behavioral and mental health outcomes, sometimes more consistently than device-based metrics.[36, 39] Moreover, sleep recommendations are largely based on self-report [40], reflecting their value for clinical and public health settings despite their limitations. Second, although the cohort was racially and ethnically diverse, subgroup sample sizes were modest for some groups, resulting in wider confidence intervals in stratified analyses; therefore, race-stratified findings should be interpreted as exploratory and hypothesis-generating rather than definitive evidence of differential effects. Third, residual confounding cannot be excluded. Despite adjustment for key sociodemographic and behavioral covariates, unmeasured factors, such as occupational schedules, environmental conditions, psychosocial stress exposure, medication use, or genetic susceptibility, common sleep disorders during pregnancy (e.g., sleep apnea, restless leg syndrome) may influence both sleep health and long-term weight outcomes, and structural determinants of sleep health are well documented.[13] Fourth, given the number of comparisons examined—multiple sleep domains across two pregnancy time points and two weight outcomes—the possibility of Type I error cannot be excluded. We did not adjust for multiple comparisons; however, this study is largely exploratory in nature, aimed at determining whether multidimensional sleep health or individual sleep domains at different pregnancy time points are associated with long-term weight outcomes and warrant further investigation. Individual findings should therefore be interpreted in the context of the overall pattern of results rather than as confirmatory evidence. Fifth, pre-pregnancy weight was self-reported at the first study visit, which may introduce error into the weight-retention outcome. Sixth, because casewise deletion produced non-identical early- and mid-pregnancy analytic samples, differences between the two time points may partly reflect differences in sample composition and precision rather than a true effect of exposure timing; this qualifies our interpretation of mid-pregnancy as a distinct window. Seventh, follow-up ranged from 2 to 7 years and participants could have had subsequent pregnancies before the follow-up visit, contributing variability to measured weight retention. Finally, participants included in the analytic sample were generally older and more socioeconomically advantaged than those excluded, which may limit generalizability to more socioeconomically vulnerable populations; however, differences in pre-pregnancy BMI and multidimensional sleep health were modest, suggesting that selection factors are unlikely to fully account for the observed associations.

## Conclusions

Multidimensional sleep health during mid-pregnancy, rather than early gestation, was modestly associated with lower weight retention 2 to 7 years postpartum. These findings support conceptualizing sleep as an integrated behavioral construct and suggest that mid-gestation may represent a sensitive window during which cumulative sleep patterns influence long-term metabolic regulation. Although effect sizes were modest, even small differences in postpartum weight retention may translate into meaningful cardiometabolic implications over the reproductive life course. Whether improving sleep duration and quality during pregnancy reduces long-term weight retention remains to be tested in intervention studies. Future research incorporating objective sleep measures, mechanistic biomarkers, and detailed assessments of contextual and structural influences will be essential for clarifying pathways and identifying optimal targets for preventive intervention.

## Data Availability

Due to the sensitive nature of the data collected in this study, requests to access data used in this analysis from qualified researchers trained in human subject confidentiality protocols will be considered by the data coordinating center and may be submitted at https://numom2b.org.

## Acknowledgement

None

## Notes

Funding source: This study was supported by grant funding from the Eunice Kennedy Shriver National Institute of Child Health and Human Development (NICHD): U10 HD063036; U10 HD063072; U10 HD063047; U10 HD063037; U10 HD063041; U10 HD063020; U10 HD063046; U10 HD063048; and U10 HD063053. In addition, support was provided by Clinical and Translational Science Institutes: UL1TR001108 and UL1TR000153. Additionally, this study was supported by Cooperative agreement funding from the National Heart, Lung, and Blood Institute and the Eunice Kennedy Shriver National Institute of Child Health and Human Development: U10 HL119991; U10 HL119989; U10 HL120034; U10 HL119990; U10 HL120006; U10 HL119992; U10 HL120019; U10-HL119993; U10 HL120018, and U01HL145358; and the National Center for Advancing Translational Sciences through UL 1 TR000124, UL 1 TR000153, UL 1 TR000439, and UL 1 TR001108; U54 AG065141, U54 AG094168 and the Barbra Streisand Women’s Cardiovascular Research and Education Program, and the Erika J. Glazer Women’s Heart Research Initiative, Cedars-Sinai Medical Center, Los Angeles.

### Competing Interest Statement

Phyllis C. Zee: Consultant and scientific advisor for Eisai, Takeda, Alkermes, NextSense

### Author Declarations

The Institutional Review Boards at all participating institutions (RTI International, Case Western Reserve University, Columbia University, Indiana University, University of Pittsburgh, Northwestern University, University of California, Irvine, University of Pennsylvania, University of Utah) approved this study, and all participants provided written informed consent prior to participation.

