## Supplemental Figure 1 for "Relations Between Prenatal Sleep Health and Maternal Weight Retention 2 to 7 Years After a First Birth: The NuMoM2b-HHS"

**Supplemental Figure 1 – Participant flow chart**

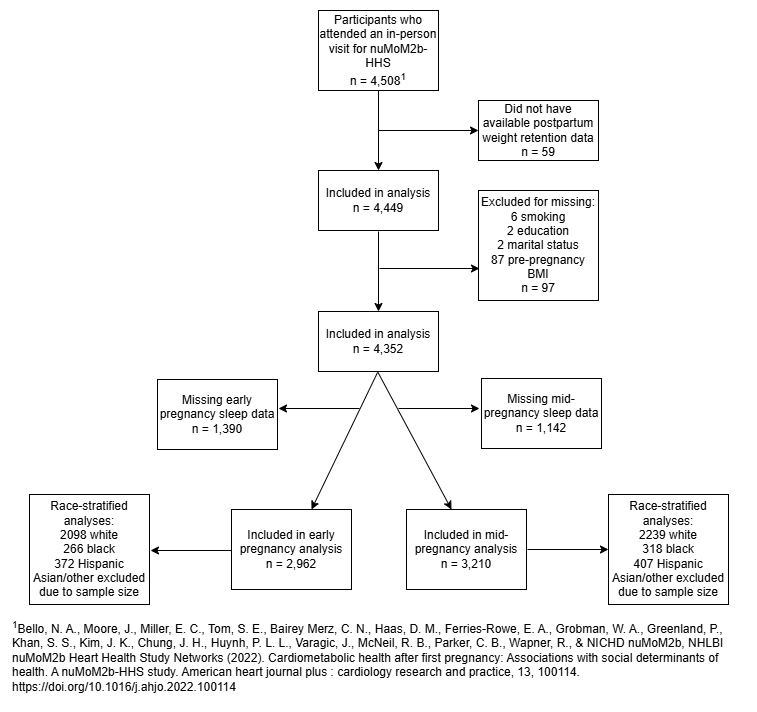

| Supplemental Table 1 - Baseline Characteristics of Participants by Inclusion Status | | | | | | |
| --- | --- | --- | --- | --- | --- | --- |
|  | Early-Pregnancy Sample | | | Mid-Pregnancy Sample | | |
| Characteristic^1^ | **Excluded**  N = 1487^1^ | **Included**  N = 2962^1^ | **p-value**^1^ | **Excluded**  N = 1239^1^ | **Included**  N = 3210^1^ | **p-value**^1^ |
| Age (years) | 24 (20, 30) | 28 (24, 32) | <0.001 | 24 (20, 29) | 28 (24, 32) | <0.001 |
| Race/Ethnicity |  |  | <0.001 |  |  | <0.001 |
| Non-Hispanic White | 672 (45%) | 2098 (71%) |  | 531 (43%) | 2239 (70%) |  |
| Non-Hispanic Black | 341 (23%) | 266 (9.0%) |  | 289 (23%) | 318 (9.9%) |  |
| Hispanic | 357 (24%) | 372 (13%) |  | 322 (26%) | 407 (13%) |  |
| Asian | 43 (2.9%) | 91 (3.1%) |  | 27 (2.2%) | 107 (3.3%) |  |
| Other | 74 (5.0%) | 135 (4.6%) |  | 70 (5.6%) | 139 (4.3%) |  |
| Education Level |  |  | <0.001 |  |  | <0.001 |
| < High School | 466 (31%) | 349 (12%) |  | 410 (33%) | 405 (13%) |  |
| High School | 351 (24%) | 531 (18%) |  | 282 (23%) | 600 (19%) |  |
| > High School | 668 (45%) | 2082 (70%) |  | 545 (44%) | 2205 (69%) |  |
| Missing | 2 | 0 |  | 2 | 0 |  |
| Income (% of Federal Poverty Level) | 230 (90, 448) | 448 (230, 640) | <0.001 | 230 (81, 448) | 435 (191, 640) | <0.001 |
| Missing | 395 | 392 |  | 348 | 439 |  |
| Pre-Pregnancy BMI | 25 (22, 30) | 24 (21, 28) | <0.001 | 25 (22, 30) | 24 (21, 28) | <0.001 |
| Missing | 94 | 0 |  | 94 | 0 |  |
| Smoked 3 Months Prior to Pregnancy | 292 (20%) | 416 (14%) | <0.001 | 226 (18%) | 482 (15%) | 0.007 |
| Missing | 6 | 0 |  | 6 | 0 |  |
| EPDS Score | 6.0 (3.0, 9.0) | 5.0 (2.0, 8.0) | <0.001 | 6.0 (3.0, 9.0) | 5.0 (2.0, 8.0) | <0.001 |
| Missing | 98 | 23 |  | 55 | 66 |  |
| High-Intensity Exercise (mins/week) | 0 (0, 0) | 0 (0, 60) | <0.001 | 0 (0, 0) | 0 (0, 60) | <0.001 |
| Missing | 6 | 0 |  | 6 | 0 |  |
| Low-Intensity Exercise (mins/week) | 0 (0, 120) | 60 (0, 140) | <0.001 | 0 (0, 120) | 50 (0, 135) | <0.001 |
| Missing | 6 | 0 |  | 6 | 0 |  |
| Marital Status |  |  | <0.001 |  |  | <0.001 |
| Not Married | 823 (56%) | 870 (29%) |  | 717 (58%) | 976 (30%) |  |
| Married | 658 (44%) | 2092 (71%) |  | 516 (42%) | 2234 (70%) |  |
| Missing | 6 | 0 |  | 6 | 0 |  |
| gravidity_v1 |  |  | 0.2 |  |  | 0.084 |
| Median (Q1, Q3) | 1.00 (1.00, 2.00) | 1.00 (1.00, 1.00) |  | 1.00 (1.00, 2.00) | 1.00 (1.00, 1.00) |  |
| Has Government Insurance | 657 (44%) | 590 (20%) | <0.001 | 576 (46%) | 671 (21%) | <0.001 |
| Has Military Insurance | 5 (0.3%) | 16 (0.5%) | 0.3 | 5 (0.4%) | 16 (0.5%) | 0.7 |
| Has Commercial Insurance | 787 (53%) | 2317 (78%) | <0.001 | 625 (50%) | 2479 (77%) | <0.001 |
| Self-Pay for Healthcare | 47 (3.2%) | 72 (2.4%) | 0.2 | 44 (3.6%) | 75 (2.3%) | 0.024 |
| Sleep Duration (hours) | 8.14 (7.33, 9.02) | 7.96 (7.25, 8.64) | <0.001 | 8.07 (7.14, 9.06) | 7.98 (7.30, 8.64) | 0.040 |
| Missing | 715 | 0 |  | 400 | 315 |  |
| Binary Sleep Duration |  |  | <0.001 |  |  | <0.001 |
| < 7 or > 9 hours | 336 (44%) | 992 (33%) |  | 395 (47%) | 933 (32%) |  |
| 7-9 hours | 436 (56%) | 1970 (67%) |  | 444 (53%) | 1962 (68%) |  |
| Missing | 715 | 0 |  | 400 | 315 |  |
| Wake After Sleep Onset | 5 (0, 20) | 15 (5, 30) | <0.001 | 10 (3, 30) | 10 (5, 30) | 0.039 |
| Missing | 192 | 0 |  | 192 | 0 |  |
| Binary Wake After Sleep Onset |  |  | <0.001 |  |  | 0.3 |
| > 40 minutes | 139 (11%) | 500 (17%) |  | 167 (16%) | 472 (15%) |  |
| ≤ 40 minutes | 1156 (89%) | 2462 (83%) |  | 880 (84%) | 2738 (85%) |  |
| Missing | 192 | 0 |  | 192 | 0 |  |
| Daytime Sleepiness | 8.0 (5.0, 11.0) | 7.0 (5.0, 10.0) | 0.2 | 8.0 (5.0, 11.0) | 7.0 (5.0, 10.0) | 0.029 |
| Missing | 948 | 0 |  | 507 | 441 |  |
| Binary Daytime Sleepiness |  |  | 0.014 |  |  | 0.077 |
| Excessive Sleepiness (≥ 10 points) | 197 (37%) | 923 (31%) |  | 254 (35%) | 866 (31%) |  |
| Normal (< 10 points) | 342 (63%) | 2039 (69%) |  | 478 (65%) | 1903 (69%) |  |
| Missing | 948 | 0 |  | 507 | 441 |  |
| Sleep Regularity (hours) | 1.00 (0.50, 1.75) | 1.13 (0.63, 1.75) | 0.067 | 1.00 (0.50, 1.75) | 1.13 (0.63, 1.75) | 0.050 |
| Missing | 1056 | 0 |  | 555 | 501 |  |
| Binary Sleep Regularity |  |  | 0.019 |  |  | 0.052 |
| ≥ 1 hour | 261 (61%) | 1964 (66%) |  | 427 (62%) | 1798 (66%) |  |
| < 1 hour | 170 (39%) | 998 (34%) |  | 257 (38%) | 911 (34%) |  |
| Missing | 1056 | 0 |  | 555 | 501 |  |
| Sleep Timing (hour) | 3.45 (2.68, 4.58) | 3.20 (2.62, 3.89) | <0.001 | 3.46 (2.73, 4.46) | 3.18 (2.60, 3.90) | <0.001 |
| Missing | 715 | 0 |  | 400 | 315 |  |
| Binary Sleep Timing |  |  | <0.001 |  |  | <0.001 |
| Outside of 2-4 AM window | 351 (45%) | 866 (29%) |  | 350 (42%) | 867 (30%) |  |
| Between 2-4 AM | 421 (55%) | 2096 (71%) |  | 489 (58%) | 2028 (70%) |  |
| Missing | 715 | 0 |  | 400 | 315 |  |
| Sleep Quality (Restfulness) |  |  | 0.045 |  |  | 0.011 |
| Very sound or restful | 70 (12%) | 356 (12%) |  | 93 (13%) | 333 (12%) |  |
| Sound or restful | 144 (26%) | 824 (28%) |  | 171 (23%) | 797 (29%) |  |
| Average quality | 215 (38%) | 1236 (42%) |  | 309 (42%) | 1142 (41%) |  |
| Restless | 114 (20%) | 474 (16%) |  | 144 (19%) | 444 (16%) |  |
| Very restless | 20 (3.6%) | 72 (2.4%) |  | 25 (3.4%) | 67 (2.4%) |  |
| Missing | 924 | 0 |  | 497 | 427 |  |
| Binary Sleep Quality |  |  | 0.003 |  |  | 0.007 |
| Self-reported as worse than average | 134 (24%) | 546 (18%) |  | 169 (23%) | 511 (18%) |  |
| Self-reported as average or better | 429 (76%) | 2416 (82%) |  | 573 (77%) | 2272 (82%) |  |
| Missing | 924 | 0 |  | 497 | 427 |  |
| Multidimensional Sleep Health (From Binary Variables) |  |  | 0.2 |  |  | <0.001 |
| Median (Q1, Q3) | 4.00 (3.00, 5.00) | 4.00 (3.00, 5.00) |  | 4.00 (3.00, 5.00) | 4.00 (3.00, 5.00) |  |
| Missing | 1449 | 0 |  | 750 | 699 |  |
| ^1^This table describes early pregnancy characteristics of participants included in/excluded from the early pregnancy and mid-pregnancy samples. | | | | | | |

| **Supplemental Table 2 - Sleep Health Characteristics at Each Timepoint** | | |
| --- | --- | --- |
|  | Early Pregnancy | Mid-Pregnancy |
| **Characteristic** | **N = 2962**^1^ | **N = 3210**^1^ |
| Sleep Duration (Hours) |  |  |
| Mean (SD) | 7.94 (1.25) | 7.78 (1.26) |
| Median (Q1, Q3) | 7.96 (7.25, 8.64) | 7.81 (7.13, 8.50) |
| Binary Sleep Duration |  |  |
| < 7 or > 9 hours | 992 (33%) | 1095 (34%) |
| 7-9 hours | 1970 (67%) | 2115 (66%) |
| Wake After Sleep Onset |  |  |
| Mean (SD) | 23 (27) | 28 (31) |
| Median (Q1, Q3) | 15 (5, 30) | 20 (10, 30) |
| Binary Wake After Sleep Onset |  |  |
| > 40 minutes | 500 (17%) | 683 (21%) |
| ≤ 40 minutes | 2462 (83%) | 2527 (79%) |
| Daytime Sleepiness |  |  |
| Mean (SD) | 7.7 (4.0) | 6.6 (4.1) |
| Median (Q1, Q3) | 7.0 (5.0, 10.0) | 6.0 (4.0, 9.0) |
| Binary Daytime Sleepiness |  |  |
| Excessive Sleepiness (≥ 10 points) | 923 (31%) | 737 (23%) |
| Normal (< 10 points) | 2039 (69%) | 2473 (77%) |
| Sleep Regularity (Hours) |  |  |
| Mean (SD) | 1.08 (1.98) | 0.90 (1.78) |
| Median (Q1, Q3) | 1.13 (0.63, 1.75) | 1.00 (0.50, 1.50) |
| Binary Sleep Regularity |  |  |
| ≥ 1 hour | 1964 (66%) | 1878 (59%) |
| < 1 hour | 998 (34%) | 1332 (41%) |
| Sleep Timing |  |  |
| Mean (SD) | 3.55 (1.99) | 3.55 (1.92) |
| Median (Q1, Q3) | 3.20 (2.62, 3.89) | 3.18 (2.61, 3.95) |
| Binary Sleep Timing |  |  |
| Outside of 2-4 AM window | 866 (29%) | 970 (30%) |
| Between 2-4 AM | 2096 (71%) | 2240 (70%) |
| Sleep Quality (Restfulness) |  |  |
| Very sound or restful | 356 (12%) | 337 (10%) |
| Sound or restful | 824 (28%) | 848 (26%) |
| Average quality | 1236 (42%) | 1329 (41%) |
| Restless | 474 (16%) | 612 (19%) |
| Very restless | 72 (2.4%) | 84 (2.6%) |
| Binary Sleep Quality |  |  |
| Self-reported as worse than average | 546 (18%) | 696 (22%) |
| Self-reported as average or better | 2416 (82%) | 2514 (78%) |
| Multidimensional Sleep Health (From Binary Variables) |  |  |
| Mean (SD) | 4.04 (1.19) | 4.11 (1.24) |
| Median (Q1, Q3) | 4.00 (3.00, 5.00) | 4.00 (3.00, 5.00) |
| Continuous Weight Change (lbs) |  |  |
| Mean (SD) | 11 (24) | 10 (23) |
| Median (Q1, Q3) | 7 (0, 19) | 7 (0, 18) |
| Substantial Weight Change (≥11 lbs) | 1194 (40%) | 1300 (40%) |
| Incident Overweight/Obesity | 394 (22%) | 418 (22%) |
| Missing | 1203 | 1305 |
| ^1^n (%) | | |
| This table describes sleep health during early and mid-pregnancy. | | |

| **Supplementary Table 3 – Evaluation of interaction by race/ethnicity on the association between pregnancy sleep health indicators and continuous postpartum weight retention (lbs).** | | | | |
| --- | --- | --- | --- | --- |
|  | **Non-Hispanic Black** | | **Hispanic** | |
| **Sleep Metric** | **Interaction Beta (95% CI)** | **p-value** | **Interaction Beta (95% CI)** | **p-value** |
| **Early Pregnancy** | | | | |
| Multidimensional Sleep Health | -0.09 (-2.45, 2.28) | 0.942 | -2.45 (-5.01, 0.12) | 0.061 |
| Sleep Timing | -0.55 (-6.87, 5.77) | 0.864 | -1.01 (-7.73, 5.71) | 0.768 |
| Sleep Duration | -0.01 (-6.17, 6.15) | 0.998 | -3.93 (-10.31, 2.45) | 0.227 |
| Sleep Regularity | 2.86 (-3.41, 9.13) | 0.371 | -1.05 (-8.15, 6.05) | 0.772 |
| Wake After Sleep Onset | 0.67 (-7.05, 8.39) | 0.865 | -6.13 (-12.49, 0.23) | 0.059 |
| Sleep Quality | -0.83 (-8.6, 6.93) | 0.833 | -0.24 (-8.07, 7.58) | 0.951 |
| Daytime Sleepiness | -4.02 (-10.46, 2.41) | 0.220 | -6.63 (-14.57, 1.3) | 0.101 |
| **Mid-pregnancy** | | | | |
| Multidimensional Sleep Health | -0.11 (-2.45, 2.23) | 0.924 | -1.13 (-3.41, 1.14) | 0.328 |
| **Sleep Timing** | **1.33 (-4.75, 7.4)** | **0.668** | **-9.38 (-15.26, -3.51)** | **0.002** |
| Sleep Duration | 0.25 (-5.49, 5.99) | 0.932 | -1.05 (-6.77, 4.67) | 0.720 |
| Sleep Regularity | -3.34 (-9.05, 2.37) | 0.252 | -1.84 (-7.47, 3.79) | 0.521 |
| Wake After Sleep Onset | -1.56 (-9.53, 6.4) | 0.701 | 1.37 (-6.04, 8.78) | 0.717 |
| **Sleep Quality** | **3.59 (-2.39, 9.56)** | **0.239** | **6.92 (0.64, 13.19)** | **0.031** |
| Daytime Sleepiness | -1.24 (-7.7, 5.23) | 0.708 | -1.36 (-8.28, 5.56) | 0.701 |
| Abbreviations: CI = Confidence Interval; NHW = Non-Hispanic White. | | | | |
| Interpretation: The Interaction Beta represents the absolute difference in the effect of poor sleep on continuous weight retention (in lbs) between the specified racial/ethnic group and the NHW reference group. A significant p-value indicates that the association between the sleep metric and weight retention differs significantly by race/ethnicity. | | | | |

| **Supplementary Table 4 – The associations between mid-pregnancy sleep health indicators and weight retention (in lbs) from pre-pregnancy to 2 to 7 years postpartum overall and by race/ethnicity.** | | | | | | | | |
| --- | --- | --- | --- | --- | --- | --- | --- | --- |
|  | Overall N = 3210 | | Non-Hispanic White N = 2239 | | Non-Hispanic Black N = 318 | | Hispanic N = 407 | |
| Sleep Domain | Beta | 95% CI^1^ | Beta | 95% CI^1^ | Beta | 95% CI^1^ | Beta | 95% CI^1^ |
| Multidimensional Sleep Health | -0.82 | -1.47, -0.17 | -0.90 | -1.66, -0.15 | -1.1 | -3.23, 1.01 | 0.14 | -2.06, 2.35 |
| Sleep Timing |  |  |  |  |  |  |  |  |
| Outside of 2-4 AM window | — | — | — | — | — | — | — | — |
| Between 2-4 AM | -0.89 | -2.71, 0.92 | -2.1 | -4.30, 0.06 | -4.4 | -9.94, 1.10 | 7.0 | 1.34, 12.59 |
| Sleep Duration |  |  |  |  |  |  |  |  |
| < 7 or > 9 hours | — | — | — | — | — | — | — | — |
| 7-9 hours | -2.4 | -4.12, -0.70 | -2.3 | -4.35, -0.33 | -3.3 | -8.78, 2.14 | -1.4 | -6.89, 4.12 |
| Sleep Regularity |  |  |  |  |  |  |  |  |
| ≥ 1 hour | — | — | — | — | — | — | — | — |
| < 1 hour | 0.98 | -0.64, 2.60 | 0.05 | -1.82, 1.93 | 3.8 | -1.63, 9.28 | 2.5 | -2.97, 7.92 |
| Wake After Sleep Onset |  |  |  |  |  |  |  |  |
| > 40 minutes | — | — | — | — | — | — | — | — |
| ≤ 40 minutes | -0.93 | -2.88, 1.02 | -0.77 | -2.97, 1.43 | 0.56 | -6.06, 7.18 | -2.2 | -9.13, 4.74 |
| Sleep Quality |  |  |  |  |  |  |  |  |
| Self-reported as worse than average | — | — | — | — | — | — | — | — |
| Self-reported as average or better | -2.4 | -4.31, -0.44 | -0.72 | -2.93, 1.49 | -4.9 | -11.31, 1.57 | -7.9 | -14.59, -1.25 |
| Daytime Sleepiness |  |  |  |  |  |  |  |  |
| Excessive Sleepiness (≥ 10 points) | — | — | — | — | — | — | — | — |
| Normal (< 10 points) | -1.1 | -3.02, 0.78 | -1.3 | -3.56, 0.96 | -0.53 | -6.26, 5.19 | -0.15 | -6.39, 6.09 |
| ^1^All models adjusted for age, education, marital status, pre-pregnancy BMI, and smoking status. Overall models also adjusted for race. Overall models included Asian and Other groups, which were excluded from race-stratified models due to sample size considerations. | | | | | | | | |

| **Supplementary Table 5 – Associations of early and mid-pregnancy sleep health indicators with substantial postpartum weight retention (≥11 lbs), including evaluation of additive and multiplicative interaction by race/ethnicity.** | | | | | | | | | |
| --- | --- | --- | --- | --- | --- | --- | --- | --- | --- |
|  | **NHW** | **Non-Hispanic Black** | | | | **Hispanic** | | | |
|  | **RR_S_** | **RR_R_** | **RR_SR_** | **RERI (95% CI)** | **Mult. Ratio (95% CI)** | **RR_R_** | **RR_SR_** | **RERI (95% CI)** | **Mult. Ratio (95% CI)** |
| **Sleep Metric** |  |  |  |  |  |  |  |  |  |
| **Early Pregnancy** | | | | | | | | | |
| **Multidimensional Sleep Health** | **1.03** | **0.84** | **0.91** | **0.04 (-0.04, 0.12)** | **1.05 (0.95, 1.17)** | **1.53** | **1.37** | **-0.19 (-0.34, -0.03)** | **0.87 (0.79, 0.96)** |
| Sleep Timing | 1.05 | 0.89 | 1.07 | 0.13 (-0.12, 0.38) | 1.15 (0.89, 1.48) | 1.25 | 1.04 | -0.26 (-0.52, 0) | 0.79 (0.63, 1) |
| Sleep Duration | 1.07 | 0.93 | 1.06 | 0.05 (-0.2, 0.3) | 1.06 (0.82, 1.36) | 1.22 | 1.11 | -0.18 (-0.44, 0.08) | 0.85 (0.68, 1.06) |
| Sleep Regularity | 0.91 | 0.97 | 0.87 | -0.01 (-0.24, 0.22) | 0.99 (0.77, 1.26) | 1.08 | 1.07 | 0.09 (-0.15, 0.33) | 1.10 (0.87, 1.39) |
| Wake After Sleep Onset | 1.02 | 0.95 | 1.07 | 0.10 (-0.22, 0.42) | 1.10 (0.81, 1.51) | 1.17 | 1.01 | -0.19 (-0.52, 0.14) | 0.84 (0.61, 1.15) |
| Sleep Quality | 1.06 | 0.94 | 1.12 | 0.12 (-0.2, 0.43) | 1.12 (0.83, 1.51) | 1.16 | 1.13 | -0.10 (-0.4, 0.21) | 0.91 (0.7, 1.2) |
| **Daytime Sleepiness** | **1.12** | **0.99** | **1.03** | **-0.08 (-0.33, 0.18)** | **0.93 (0.73, 1.19)** | **1.29** | **0.92** | **-0.48 (-0.78, -0.19)** | **0.64 (0.48, 0.85)** |
| **Mid-pregnancy** | | | | | | | | | |
| Multidimensional Sleep Health | 1.06 | 0.99 | 1.04 | -0.01 (-0.09, 0.07) | 0.99 (0.91, 1.08) | 1.29 | 1.27 | -0.08 (-0.18, 0.03) | 0.93 (0.86, 1.01) |
| **Sleep Timing** | **1.06** | **0.91** | **1.11** | **0.13 (-0.1, 0.36)** | **1.14 (0.91, 1.43)** | **1.26** | **0.99** | **-0.34 (-0.59, -0.08)** | **0.74 (0.59, 0.92)** |
| Sleep Duration | 1.11 | 1.00 | 1.06 | -0.04 (-0.28, 0.19) | 0.96 (0.77, 1.21) | 1.16 | 1.17 | -0.10 (-0.35, 0.15) | 0.91 (0.73, 1.13) |
| Sleep Regularity | 1.01 | 1.08 | 0.91 | -0.18 (-0.41, 0.05) | 0.83 (0.66, 1.04) | 1.18 | 1.07 | -0.12 (-0.36, 0.12) | 0.90 (0.72, 1.12) |
| Wake After Sleep Onset | 1.10 | 1.00 | 1.07 | -0.03 (-0.31, 0.25) | 0.97 (0.75, 1.27) | 1.15 | 1.14 | -0.11 (-0.42, 0.21) | 0.90 (0.68, 1.19) |
| **Sleep Quality** | **1.05** | **0.96** | **1.12** | **0.11 (-0.15, 0.37)** | **1.11 (0.87, 1.42)** | **1.05** | **1.45** | **0.36 (0.08, 0.64)** | **1.32 (1.05, 1.66)** |
| Daytime Sleepiness | 1.13 | 0.98 | 1.07 | -0.04 (-0.29, 0.21) | 0.96 (0.76, 1.22) | 1.15 | 1.14 | -0.14 (-0.43, 0.15) | 0.87 (0.68, 1.13) |
| Abbreviations: CI = Confidence Interval; NHW = Non-Hispanic White; RERI = Relative Excess Risk due to Interaction; RR = Relative Risk. | | | | | | | | | |
| Column Explanations: RR_S_ = The relative risk of substantial weight retention associated with poor sleep health among Non-Hispanic White participants; RR_R_ = The relative risk among the specified racial/ethnic group with favorable sleep health compared to NHW with favorable sleep health; RR_SR_ = The relative risk among the specified racial/ethnic group with poor sleep health compared to NHW with favorable sleep health. | | | | | | | | | |
| Interaction Calculations: Additive interaction (RERI) was calculated as RR_SR_ - RR_S_ - RR_R_ + 1. Multiplicative interaction (Mult. Ratio) was calculated as the ratio of the joint effect to the product of the independent effects: RR_SR_ / (RR_S_ * RR_R_). | | | | | | | | | |

| **Supplementary Table 6 – The associations between early pregnancy sleep health indicators and substantial weight retention (≥11 lbs) from pre-pregnancy to 2 to 7 years postpartum overall and by race/ethnicity.** | | | | | | | | |
| --- | --- | --- | --- | --- | --- | --- | --- | --- |
|  | Overall N = 2962 | | Non-Hispanic White N = 2098 | | Non-Hispanic Black N = 266 | | Hispanic N = 372 | |
| Sleep Domain | Risk Ratio | 95% CI^1^ | Risk Ratio | 95% CI^1^ | Risk Ratio | 95% CI^1^ | Risk Ratio | 95% CI^1^ |
| Multidimensional Sleep Health | 0.99 | 0.95, 1.02 | 0.98 | 0.94, 1.03 | 0.92 | 0.83, 1.01 | 1.1 | 1.02, 1.21 |
| Sleep Timing |  |  |  |  |  |  |  |  |
| Outside of 2-4 AM window | — | — | — | — | — | — | — | — |
| Between 2-4 AM | 0.99 | 0.90, 1.08 | 0.98 | 0.87, 1.12 | 0.81 | 0.64, 1.02 | 1.2 | 0.96, 1.45 |
| Sleep Duration |  |  |  |  |  |  |  |  |
| < 7 or > 9 hours | — | — | — | — | — | — | — | — |
| 7-9 hours | 0.97 | 0.88, 1.06 | 0.95 | 0.85, 1.07 | 0.88 | 0.71, 1.10 | 1.1 | 0.87, 1.32 |
| Sleep Regularity |  |  |  |  |  |  |  |  |
| ≥ 1 hour | — | — | — | — | — | — | — | — |
| < 1 hour | 1.1 | 0.98, 1.17 | 1.1 | 0.98, 1.23 | 1.1 | 0.88, 1.38 | 1.1 | 0.86, 1.31 |
| Wake After Sleep Onset |  |  |  |  |  |  |  |  |
| > 40 minutes | — | — | — | — | — | — | — | — |
| ≤ 40 minutes | 0.98 | 0.87, 1.09 | 0.98 | 0.86, 1.13 | 0.89 | 0.68, 1.18 | 1.1 | 0.83, 1.48 |
| Sleep Quality |  |  |  |  |  |  |  |  |
| Self-reported as worse than average | — | — | — | — | — | — | — | — |
| Self-reported as average or better | 0.94 | 0.84, 1.04 | 0.96 | 0.84, 1.09 | 0.85 | 0.65, 1.12 | 0.99 | 0.78, 1.26 |
| Daytime Sleepiness |  |  |  |  |  |  |  |  |
| Excessive Sleepiness (≥ 10 points) | — | — | — | — | — | — | — | — |
| Normal (< 10 points) | 0.97 | 0.88, 1.06 | 0.90 | 0.80, 1.01 | 0.95 | 0.76, 1.19 | 1.4 | 1.09, 1.83 |
| ^1^All models adjusted for age, education, marital status, pre-pregnancy BMI, and smoking status. Overall models also adjusted for race. Overall models included Asian and Other groups, which were excluded from race-stratified models due to sample size considerations. | | | | | | | | |
